# Trends in Pre-Exposure Prophylaxis Uptake Among Adolescent Girls and Young Women in Gauteng Province, South Africa: A Study Protocol

**DOI:** 10.1101/2025.09.29.25336917

**Authors:** Salome Thilivhali Sigida, Lufuno Makhado, Thendo Gertie Makhado

**Author notes:** Funding: None. Competing Interests: The authors declare no competing interests. Data Availability: Data are available from the corresponding author upon reasonable request.

## Abstract

Oral pre-exposure prophylaxis (PrEP) plays an important role as part of a combination HIV prevention strategy for adolescent girls and young women (AGYW), a sub-population at continued elevated risk of HIV, compared to their male peers. The current study aims to analyze temporal trends and demographic patterns in PrEP uptake among AGYW aged 15–24 in Gauteng Province, focusing specifically on Tshwane and Ekurhuleni Districts. The quantitative approach, specifically a longitudinal retrospective cohort design, will be used to examine the temporal trends and demographic patterns of PrEP uptake amongst the AGYWs in Gauteng. The study will be conducted within a post-positivist epistemological approach using secondary data to investigate trends and patterns in PrEP use, particularly among AGYWs aged 15–24 years in Gauteng Province, primarily in Tshwane and Ekurhuleni District. The study will employ a total population sampling of all who meet the inclusion criteria. The data from the current study will be obtained from the DHIS electronic database. Data management and analysis will be performed using SPSS, ensuring a robust and systematic examination of the data. The coding procedures, protocols for data cleaning, and statistical techniques to minimize errors and ensure consistent analysis will all further support reliability. The construct validity will be ensured by showing how the findings from each included dataset reflect the constructs they are studying, such as PrEP use, adherence, and HIV incidence among AGYW. Ethical considerations will be ensured, and results will be interpreted according to the findings of the study

## Introduction

In 2023, it was estimated that some 3800 adolescent girls and young women (AGYW) acquired HIV infection weekly, with 76% of the new infections occurring in sub-Saharan Africa (SSA) (UNAIDS, 2024). In sub-Saharan Africa (SSA) in 2023, 198 000 AGYW (15–24 years) were considered newly infected with HIV and were eight times more vulnerable to acquired HIV as their male counterparts (UNAIDS, 2024). The high burden of HIV among AGYW in the region is fuelled by numerous biological, social, and structural drivers, including gender-based violence, limited access to care including sexual and reproductive health (SRH) care, and economic instability (Chen et al, 2025).

South Africa bears the world’s largest HIV burden, with an estimated 7.8 million of its citizens currently living with HIV (Payagala & Pozniak, 2024). The findings of the HSRC survey indicated that the national estimate of HIV prevalence in South Africa for all ages in 2022 was 12.7%, down from 14% in 2017. HIV prevalence in the 25–49 age group has decreased significantly between the two surveys (Payagala & Pozniak, 2024).

Substantial progress has been made in the treatment of HIV in South Africa, where an extensive antiretroviral therapy (ART) program now supports over 5.5 million individuals. Despite these advancements, the country continues to grapple with a high HIV prevalence, as outlined by Nyasulu et al. (2016). The HIV/AIDS epidemic reveals significant demographic and regional disparities, with specific populations and geographic areas experiencing a disproportionately greater impact (Brawner et al., 2017).

Zuma et al. (2022) identify various socio-behavioral and structural factors that drive the HIV epidemic. These factors include multiple sexual partnerships, unprotected sexual intercourse, inconsistent condom use, socioeconomic inequalities, harmful gender norms, sexual violence, and the consumption of alcohol and other substances. In addition, Madiba and Ngwenya (2017) highlight that specific socio-behavioral characteristics, such as the age of sexual debut among youth and the presence of age-disparate relationships, are linked to increased vulnerability to HIV.

Notably, age-disparate sexual partnerships, particularly those involving young women and older men, have been instrumental in sustaining high HIV prevalence rates within the 15–24 age group (Madiba & Ngwenya, 2017). This demographic is particularly at risk, facing an eight-fold greater likelihood of HIV infection compared to their male counterparts. Addressing these intertwined factors is critical for enhancing the effectiveness of HIV prevention and treatment strategies in South Africa.

The gender gap that emerges during adolescence and young adulthood is significantly influenced by biological, behavioral, and structural vulnerabilities. In South Africa, approximately 1,200 new HIV infections occur each week among adolescent girls and young women (AGYWs), accounting for about 25% of all new infections in the country, despite this group representing only 10% of the population (Payagala & Pozniak, 2024). Various risk factors contribute to women’s increased vulnerability to HIV acquisition, including biological susceptibility, age-disparate relationships, limited economic opportunities, gender-based violence, inadequate access to healthcare, and persistent gender inequities (Shisana et al., 2012; Dellar, Dlamini, & Karim, 2015; Machur et al., 2022). These elements create a complex risk landscape that traditional prevention strategies have inadequately addressed.

A pivotal advancement in HIV prevention is pre-exposure prophylaxis (PrEP), which involves the use of antiretroviral medications by HIV-negative individuals to reduce the risk of HIV acquisition. Daily oral PrEP, specifically with the combination of tenofovir disoproxil fumarate and emtricitabine (TDF-FTC), has been shown to reduce the risk of HIV infection by over 90% when taken consistently, as evidenced by randomized controlled trials (Fonner et al., 2016). Its high efficacy in specific populations, coupled with an acceptable safety profile, positions PrEP as a potentially transformative tool in the HIV prevention arsenal, particularly for high-incidence populations that may have limited control over traditional prevention methods.

In this context, South Africa has emerged as a leader in the clinical implementation of PrEP within sub-Saharan Africa. The country approved the use of oral PrEP in 2015 and initiated its first demonstration projects targeting key populations, including female sex workers, in urban centres (Milimu et al, 2024). Acknowledging the disproportionate impact of HIV on young people, the National Department of Health began expanding access to PrEP for university students through campus health services in 2016. In light of emerging evidence and persistent advocacy efforts, national guidelines developed in 2017-2018 explicitly recommended PrEP for AGYWs at substantial risk of HIV infection, based on a comprehensive set of clinical and behavioral criteria (Milimu et al, 2024). This coordinated effort illustrates a strategic response to the unique vulnerabilities of AGYWs and highlights the importance of targeted prevention initiatives in addressing the HIV epidemic effectively.

The National Strategic Plan for HIV, TB, and STIs (2017-2022) reaffirmed the commitment to addressing these public health challenges within the framework of the SBCC policy. Central to this plan is the inclusion of pre-exposure prophylaxis (PrEP) as a key prevention strategy and the establishment of ambitious targets aimed explicitly at adolescent girls and young women (AGYW). A comprehensive policy framework was developed to ensure that at least 85,000 AGYW receive PrEP through a variety of service delivery platforms, including primary healthcare facilities (PHCs), youth-friendly services (YFSs), and community-based approaches (South African National AIDS Council [SANAC], 2017).

Despite these progressive policy advancements and the apparent prioritization of PrEP in national health agendas, implementing PrEP programs for AGYW in South Africa has encountered significant barriers throughout the prevention cascade. These challenges span the entire continuum, from creating awareness and generating demand to facilitating initiation and ensuring the continuation of PrEP use.

In light of these obstacles, this study aims to investigate the trends in PrEP uptake among adolescent girls and young women in South Africa. By examining these trends, the research seeks to identify factors that influence PrEP utilization and to inform strategies for improving access and adherence to PrEP among this vulnerable population.

This study seeks to fill a critical evidence gap by identifying demographic and geographic trends in PrEP uptake, supporting tailored interventions to enhance reach and efficacy.

### Conceptual Framework: The Prevention’s Socio-Ecological Model

In order to understand the trends in PrEP uptake amongst AGYW in Gauteng, the present study will apply the Socio-Ecological model. The social-ecological model (SEM) is a theory of health that recognises and describes multilevel factors influencing AGYW in Gauteng to take PrEP as a HIV prevention method (Bronfenbrenner, 1979; McLeroy et al., 1988). This model acknowledges that health behaviours, including choices about HIV prevention, are influenced by overlapping spheres of individual characteristics and broad societal factors. For AGYWs in South Africa, a population already disproportionately affected by HIV with AGYW infection rates 2-3 times higher than that of male peers, examining PrEP take up calls for consideration of the nuanced interplay across multiple ecological levels, beyond even that of individual knowledge or risk perception (Celum et al., 2019; Martin et al., 2025). The SEM supports the systematized examination of entry points, barriers and facilitators, with respect to each level, including individual knowledge and risk perception, and structural determinants, such as healthcare access and gendered power structures (Zuma et al, 2022). This framework has been used in other HIV prevention studies to contextualise facilitators and barriers to uptake of prevention methods, use, and service delivery (Kaufman et al., 2014). The SEM is made up of four constructs, each representing a level of influence namely: (i) individual factors such as knowledge, attitudes, and personal beliefs (ii) interpersonal/relationship, such as the influence of partners, family and peers;

1. health system, such as service delivery models and provider attitudes and (iv) community/societal factors such as social norms and community beliefs or myths (Kayesu et al., 2022; Muhumuz et al., 2021).

## Materials and Methods Study aim

This study aims to examine the temporal trends and demographic patterns of PrEP uptake among adolescent girls and young women (AGYW) aged 15–24 years in Gauteng Province, South Africa.

### Study objectives

- To analyze temporal trends in PrEP uptake among AGYW in Tshwane and Ekurhuleni Districts from January 2023 to December 2024.
- To compare PrEP uptake rates between AGYW in Tshwane and Ekurhuleni, identifying district-specific disparities.
- To determine demographic predictors (e.g., age, education level, socioeconomic status) associated with PrEP uptake among AGYW.
- To determine factors influencing adherence to PrEP use in AGYW.

### Study design and setting

The current study will use a quantitative retrospective study with secondary data analysis to assess the temporal trend of PrEP uptake in adolescents and young women (AGYW) in Gauteng. This approach makes sense for the research objectives being addressed because it enables a comprehensive exploration of longitudinal trends across multiple geographic regions and implementation contexts without the resource and time constraints of primary data collection (Tripathy, 2013).

The study will be conducted with a post-positivist epistemological approach (Ryan, 2018; Botma et al., 2022). The post-positivist epistemological approach is suitable for this current study because it acknowledges that while objective reality exists, our understanding of it is inherently imperfect and probabilistic (Phillips & Burbules, 2000). This aligns well with the current study’s aim to identify temporal trends in PrEP uptake, recognizing that multiple factors influence these patterns that cannot all be controlled or measured with absolute precision.

### Setting of the study

The data for the current study will be collected in two districts in Gauteng Province, specifically in Tshwane and Ekurhuleni districts. The Ekurhuleni and Tshwane districts are densely populated in the Gauteng Province. Both districts are urbanised, have a diverse economy, and have large urban townships.

### Study population

The study population for the current study will include records of HIV negative AGYW aged 15-24 years who were initiated into the HIV prevention programme from January 2023 to December 2024. The study site will be in Tshwane and Ekurhuleni. The sub-districts varied by setting (urban and peri-urban), socioeconomic status (indicating a vast contrast between poverty and wealth), and population density. Sites were selected based on existing relationships for program delivery and included sites frequented by AGYW (second chance matric centres, higher learning institutions, and community skills and youth centres).

### Sampling methods and sample size determination

Total population sampling will be utilized to select the HIV negative AGYWS records who enrolled in the HIV prevention programme from January 2023 to December 2024. The selected records will include all HIV negative sexually active women, aged 15-24 years, accessing PrEP and/or SRH services in Tshwane and Ekurhuleni Districts. Thus, all HIV negative AGYWs aged 15-24 years found within the sampled records will be automatically included in the study.

### Data collection

The data from the current study will be obtained from the District Health Information System (DHIS) electronic database. The DHIS uses a biomedical form to capture beneficiary data, including written beneficiary informed consent obtained in person, sociodemographic details, an HIV and STI screening test report, and a PrEP screening and initiation report. Upon receiving ethical clearance from the University of Venda and authorisation from DHIS, a de-identified dataset containing eligible participants, thus, AGYW aged 15–24 years who tested HIV-negative and were enrolled in the PrEP program, will be extracted from DHIS. The dataset will include sociodemographic information, HIV/STI screening results, PrEP eligibility assessments, and initiation records, all captured using standardized biomedical forms during routine service delivery.

### Data management plans

Data for the current study will be received on an Excel spreadsheet from DHIS. A password-protected file will then be created. Data will only be accessed by the researcher and the supervisors. Data will be imported into SPSS. Data will be labelled, and duplicates will be removed.

### Data Analysis

The data analysis will adopt a comprehensive approach, incorporating descriptive statistics, time series analysis, and inferential statistics to achieve the research objectives effectively:

- Temporal trend analysis involves analyzing how patterns evolve over time. Data will be organized by month to examine PrEP uptake over time among AGYW in Gauteng (2023– 2024) using the PrEP initiation date. A number of initiations will be aggregated per time unit and stratified by Tshwane vs. Ekurhuleni.
- Geographic pattern analysis focuses on discovering spatial distributions and trends. Local information data, such as district or GPS coordinates, will be included to analyze geographic patterns of PrEP uptake among AGYW in Gauteng. Each PrEP initiation will be mapped according to Tshwane or Ekurhuleni District.
- Meanwhile, multivariate analysis explores the various factors that influence uptake patterns. Data management and analysis will be performed using SPSS, ensuring a robust and systematic examination of the data.

### Validity and Reliability

This study will utilize data from DHIS to ensure the reliability. The contributions of these sources come from standardized data collection protocols, rigorous quality assurance processes, and consistency across time, improving the reliability and replicability of the results. The coding procedures, protocols for data cleaning, and statistical techniques to minimize errors and ensure consistent analysis will all further support reliability.

The construct validity will be ensured by showing how the findings from each included dataset reflect the constructs they are studying, such as PrEP use, adherence, and HIV incidence among AGYW. Selection of indicators in accordance with accepted definitions and measures will enhance construct validity. Using appropriate statistical controls to minimize the risk of confounders will allow internal validity to be further established through careful selection of variables. Triangulation of findings across multiple datasets will be employed to validate trends and conclusions wherever possible. To ensure external validity, the generalizability of results, the study will use large, diverse samples representative of the broader population of adolescent girls and young women in Gauteng.

### Ethical approval

In this study, the researcher will take several important steps to uphold ethical standards. First, the researcher sought ethical clearance from the University of Venda’s Human and Clinical Trial Research Ethics Committee (FHS/25/Ph/14/0508). Approval is necessary to ensure that our secondary data analysis meets ethical guidelines. Additionally, we will set up agreements with DHIS regarding data use. These agreements will outline the necessary confidentiality measures and the permissions we need for our analysis. To protect the privacy of individuals, we will analyze the data in an aggregate form. This means we will group the data so that no individual can be identified from the results. We will also present our findings at an aggregated level to maintain this anonymity. The study will comply with the South African Protection of Personal Information Act (POPIA) and international data security standards to ensure that we handle all information responsibly and securely.

### Study Status and Timeline

The study protocol had been approved by the University of Venda Higher Degrees Committee. The Protocol has been reviewed and approved by the University of Venda Ethics Clearance Committee. Data cleaning and Analysis will commence from October to December 2025. The write-up process will commence beginning of December and the mini dissertation will be submitted in January 2026.

## Discussion

South Africa continues to have a substantial disease burden attributable to HIV among AGYW (UNAIDS, 2023). Oral PrEP for HIV prevention provides effective protection against HIV and is widely available in South Africa, yet challenges persist with daily use, particularly for AGYW (Bekker & Hillier, 2022). Two next-generation long-acting PrEP options have recently been approved for use in South Africa. These modalities will contribute to a prevention landscape in which users have a choice among prevention options and can select the option that best meets their needs (Bekker & Hillier, 2022). The current study aims to analyse temporal trends and demographic patterns in PrEP uptake among AGYW aged 15–24 in Gauteng Province, focusing specifically on Tshwane and Ekurhuleni Districts. Despite efforts to scale youth-friendly SRHR services globally, barriers to accessing care for AGYW remain. Studies have shown that demographic factors such as age, education level, and socioeconomic status significantly influence PrEP uptake (Shame et al, 2021. Thus, the current study will also determine demographic predictors (e.g., age, education level, socioeconomic status) associated with PrEP uptake among AGYW.

The study limitation includes the use of secondary data, as it may limit the depth and breadth of analysis. Additionally, discrepancies in data collection methods, reporting practices, and frameworks among different regions or organizations can affect the reliability and applicability of the findings. Results will be shared with the Department of Health (Tshwane and Ekurhuleni Districts) DHIS stakeholders, published in peer-reviewed journals, and presented at relevant conferences. Any amendments to the study will be made through the university’s higher degree committee and the Human and Clinical Trial Research Ethics Committee. Only after approval, the amendment will be applied to the current study.

## Conclusions

This research study is significant because it aims to improve understanding of HIV prevention strategies, particularly regarding the uptake of PrEP among adolescent girls and young women (AGYW) in Gauteng. By highlighting disparities in PrEP coverage across different districts, the study seeks to pinpoint areas with high and low coverage, allowing for targeted resource allocation and intervention efforts. Analysing the specific barriers and facilitators to PrEP uptake in various districts may provide insights that can help design tailored interventions, considering each area’s unique socio-cultural and economic contexts. Additionally, the study will investigate demographic factors such as education level, socioeconomic status, and age variations within the AGYW category, illuminating how structural inequalities affect health outcomes. Furthermore, this research aims to enhance understanding of AGYW’s adherence to oral PrEP, as consistent use is vital for its effectiveness. Identifying underserved sub-groups may inform the development of focused interventions to meet the needs of vulnerable populations and reduce HIV incidence. The findings could significantly influence health policies and practices, offering evidence-based recommendations that are inclusive and responsive to the needs of AGYW. This study not only contributes to local and national HIV prevention strategies but also adds to the global conversation on tackling the HIV epidemic, particularly in sub-Saharan Africa, where AGYW is a key demographic at risk. Ultimately, by addressing barriers to PrEP uptake and adherence, this research supports broader public health efforts aimed at reducing HIV transmission rates in the region.

## Data Availability

No datasets were generated or analysed during the current study. All relevant data from this study will be made available upon study completion.

## Authors’ Contributions

S.T Sigida: Conceptualization, methodology and original draft writing. T.G. Makhado: Supervision and methodology. L. Makhado: Conceptualization, supervision, methodology, review, editing, and ethical compliance. All authors reviewed, edited, and approved the final protocol manuscript.

## Acknowledgements

**N/A**

## Notes

### Competing Interest Statement

The authors have declared no competing interest.

### Funding Statement

The author(s) received no specific funding for this work.

### Author Declarations

University of Venda Ressearch Ethics

